# Bayesian Network Analysis of Covid-19 data reveals higher Infection Prevalence Rates and lower Fatality Rates than widely reported

**DOI:** 10.1101/2020.05.25.20112466

**Authors:** Martin Neil, Norman Fenton, Magda Osman, Scott McLachlan

## Abstract

Widely reported statistics on Covid-19 across the globe fail to take account of both the uncertainty of the data and possible explanations for this uncertainty. In this paper we use a Bayesian Network (BN) model to estimate the Covid-19 *infection prevalence rate (IPR)* and *infection fatality rate (IFR)* for different countries and regions, where relevant data are available. This combines multiple sources of data in a single model. The results show that Chelsea Mass. USA and Gangelt Germany have relatively higher infection prevalence rates *(IPR)* than Santa Clara USA, Kobe, Japan and England and Wales. In all cases the infection prevalence is significantly higher than what has been widely reported, with much higher community infection rates in all locations. For Santa Clara and Chelsea, both in the USA, the most likely *IFR* values are 0.3-0.4%. Kobe, Japan is very unusual in comparison with the others with values an order of magnitude less than the others at, 0.001%. The *IFR* for Spain is centred around 1%. England and Wales lie between Spain and the USA/German values with an *IFR* around 0.8%. There remains some uncertainty around these estimates but an *IFR* greater than 1% looks remote for all regions/countries. We use a Bayesian technique called ‘virtual evidence’ to test the sensitivity of the *IFR* to two significant sources of uncertainty: survey quality and uncertainty about Covid-19 death counts. In response the adjusted estimates for *IFR* are most likely to be in the range 0.3%-0.5%.

## 1. Introduction

Widely reported statistics on Covid-19 across the globe fail to take account of both the uncertainty of the data and possible explanations for this uncertainty (Fenton et al, 2020; Fenton et al, 2020b). In this paper we use a Bayesian Network (BN) model to estimate the Covid-19 *infection prevalence rate (IPR)* and *infection fatality rate (IFR)* for different countries and regions, where relevant data are available. Unlike other statistical techniques that have been used to interpret Covid-19 data, BNs combine multiple sources of data in a single model that provides statistical estimates that better reflect the uncertainty regarding mechanisms that generate the data, and the amount and type of data that is available.

The paper examines results from recent serological antibody surveys carried out globally. Plainly, any uncertainty about the accuracy of serological testing will influence the estimate of the size of community infected with Covid-19 and this will in turn influence any estimate of the infection fatality rate. A serological test with low accuracy will tend to poorly estimate the size of the community infected and this, in turn, will lead to a poor estimate of the fatality rate. Also, if the fatality count is itself unreliable the fatality rate will suffer again, because if we do not know how many had the disease and how many have died, we cannot estimate the fatality rate with confidence.

The Bayesian approach is generally recognised as more advanced than classical statistical approaches that are typically applied and can therefore address more complex questions. Unfortunately, while it may seem like there has been a deluge of Covid-19 data, there is a dearth of publicly available data of the type necessary for conducting an unbiased analysis of true infection and death rates. Typically, there is also a lack of transparency surrounding the analysis and use of data, with much of the detail and data remaining secret. Hence, we have gathered data from academic papers (mainly pre-prints), press interviews, and other sources including state archives and the mass media.

The BN model (which is implemented in a commercial, state of the art, probabilistic modelling software application – AgenaRisk [Agena 2020]) provides answers to these questions, even in the presence of such basic uncertainties. Specifically:

- What is the accuracy of serological antibody testing under development and how well does it estimate population infection prevalence?
- What serological testing surveys have been done and what does the data tell us about the prevalence of community infection in different locations.
- If the serological testing surveys are imperfect what effect does this have on our estimated infection prevalence rates *(IPR)?*
- Given our prevalence estimates, how does this compare to the case infection numbers? (those who test positive for Covid-19) i.e. how much higher is actual community infection compared to reported infection?
- From reported fatality statistics what is the infection fatality rate *(IFR)?* How does serological test quality affect the reliability of these estimates? Likewise, how does uncertainty about fatality counts affect these estimates?
- How do our estimates of covid-19 *IFR* compare to influenza *IFR?*

The paper is structured as follows: In Section 2 we discuss Covid-19 testing with a particular focus on serological tests and their accuracy (sensitivity and specificity). In Section 3 we present the data and assumptions used in our analysis. In Section 4 we present the single BN model that provides answers to a series of epidemiological questions about the disease, prevalence and infection, simultaneously, and in such a way that uncertainties about the answers to one question will influence our uncertainties about the other and vice versa. The results are presented in Section 5. Our conclusions are presented in Section 6.

## 2. Covid-19 Serological Testing

What is the accuracy of serological antibody tests under development how well does it estimate population infection prevalence?

The Covid-19 *IPR* rate can be deduced from serological antibody testing, which identifies those in the tested population whose body retains an immune response arising from prior infection with Covid-19. As the disease spreads through the population, the proportion of people who develop antibodies increases and the proportion with antibodies depends on exposure and time since the disease was introduced into the community. Hence, different countries and regions will be at different points in this process and will exhibit different *IPR* rates.

The Covid-19 *IPR* rate will also be crucially dependant on our ability to accurately measure whether an individual has antibodies. Typically, serological tests of blood or other body fluids are used to detect antibody levels and hence determine either infection (IgM antibodies) or potential immunity (IgG antibodies). Presence of the latter identifies whether patients have previously had the disease, independent of whether they exhibited symptoms. Leaving aside concerns about whether immunity is temporary or indeed absent, our uncertainty about whether a patient has had Covid-19 depends on the accuracy of the serological testing process. A patient who has tested positive using a Covid-19 serological test can do so for two reasons – they genuinely have antibodies at detectable levels (these are called ‘true positives’), or the testing process falsely identifies them as antibody positive (these are called ‘false positives’). Hence, accuracy of the test is determined by its sensitivity^1^ and specificity, where:

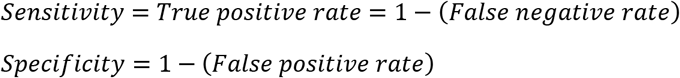

As false positive and false negative rates rise, the greater our uncertainty will be about any diagnosis of Covid-19 for an individual and across a population.

Serological testing processes, kits and machines are presently being assessed for accuracy using samples drawn from patients who are known to have the disease or are disease free. Pharmaceutical companies and academic researchers use these known samples to determine the sensitivity and specificity. At the time of writing a number of pharmaceutical serological tests have been made available in the marketplace and indeed some have received FDA emergency authorised approval for use [FDA 2020]. Accuracy for many of these tests is presently being debated.

The Covid-19 *IFR* rate is simply the number of fatalities up to some point in time divided by the size of the community infected with the virus. The number of infected will include those who test positive for antibodies, which includes those who tested positive (IgM antibodies) and those who currently test negative, but show an immune response (IgG antibodies) identifying that they were infected with Covid-19 sometime in the past. Likewise, the number of community infected cases will include those who are asymptomatic and symptomatic, whether hospitalized or not, those who finally die with (or of) the disease, and those who were infected but have fully recovered. We can estimate the number of community infected from the serological testing, and as we have said, this depends on the accuracy of the testing process. If the serological tests are inaccurate estimates of the number of infected will be more uncertain, leading to less reliable estimates for *IFR*. Similarly, the number of fatalities, and any uncertainty about the true number of fatalities, also influences the *IFR*. For example, overestimation of causalities by certifying Covid-19 deaths where patients died as a result of some pre-existing condition while infected with Covid-19, rather than only those where Covid-19 was the direct cause of their death [Fenton et al 2020] will lead to greater uncertainty about the fatality rate.

## 3. Data and Assumptions

What serological testing surveys have been done and what does the data tell us about the prevalence of community infection in different locations?

We use serological antibody survey data from these sources:

- Santa Clara County, California, USA [Bendavid et al 2020]
- Kobe, Japan [Doi 2020]
- Gangelt, Germany [Streek et al 2020]
- Chelsea, Massachusetts, USA [Saltzman 2020]
- NHS England and Wales serological survey, UK [Blanchard 2020]
- Spain [Carlos 2020]

We have also gathered and used publicly-available data on serological antibody test accuracy for each of these test sources from the FDA [FDA 2020], manufacturer websites and research papers:

- Roche, as hypothetical for England and Wales UK [FDA 2020]
- Kurabo Inc, for Kobe, Japan [Karubo 2020]
- Premier Biotech, for Santa Clara, California, USA [Bendavid et al 2020]
- EUROIMMUN for Gangelt [FDA 2020]
- BioMedomics for Chelsea, Massachusetts, USA [BioMedomics 2020]
- Orient Gene Biotech, for Spain [Carlos 2020]

It is important note that the sensitivity and specificity values are normally calculated using simply arithmetic ratios; so if the sample tested had zero false positives or false negatives then the sensitivity and specificity would be reported to be perfect (100%), no matter how many were tested. However, if you run one, and only one, test and confirm it positive, would you believe the test to be perfect on such a small experiment? This issue is addressed by the BN model. Some tests identify early or existing infection through detection of IgM antibodies. Other tests can identify patients who at some point in the past were infected with and successfully fought off the disease through detection of IgG antibodies. Many antibody tests are developed that can detect both, and where a test has this capability the regulated clinical trial and test validation process generally identifies different sensitivity and specificity values for that test for each antibody. Many of the Covid-19 antibody tests receiving emergency clearance can detect two or three antibodies but are only reporting one sensitivity and specificity value. More often this is for IgM antibodies, creating further uncertainty when we use that test to identify those who previously had the disease.

To illustrate our uncertainties about the results of serological testing we look at the false positive and false negative rates for the various serological test sources, used to estimate population infection prevalence, as shown in Table 1.

**Table 1:**
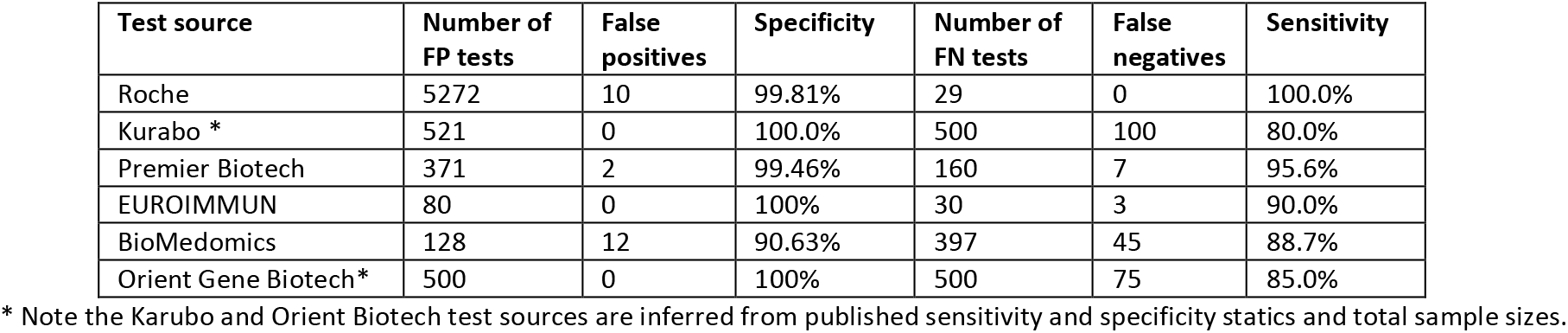
Sensitivity and specificity tests

| Test source | Number of FP tests | False positives | Specificity | Number of FN tests | False negatives | Sensitivity |
| --- | --- | --- | --- | --- | --- | --- |
| Roche | 5272 | 10 | 99.81% | 29 | 0 | 100.0% |
| Kurabo * | 521 | 0 | 100.0% | 500 | 100 | 80.0% |
| Premier Biotech | 371 | 2 | 99.46% | 160 | 7 | 95.6% |
| EUROIMMUN | 80 | 0 | 100% | 30 | 3 | 90.0% |
| BioMedomics | 128 | 12 | 90.63% | 397 | 45 | 88.7% |
| Orient Gene Biotech* | 500 | 0 | 100% | 500 | 75 | 85.0% |

\* Note the Karubo and Orient Biotech test sources are inferred from published sensitivity and specificity statics and total sample sizes.

UK fatality data and Covid-19 infection rates were collected from the UK Office for National Statistics (ONS) [ONS 2020a] and from the worldometers website [Worldometers 2020].

All available data are taken from the mid-April to early May 2020 snapshot. Thus, different countries and regions within countries will have different infection prevalence rates as these will be a determined by when the virus was first introduced and the time elapsed. For data we assume the time from infection to recovery (or fatality) is sufficient to ensure that any anyone infected will have detectable traces of antibodies in their blood.

## 4. The Bayesian Network (BN) Model

A BN is a graphical model consisting of nodes and arcs where the nodes represent variables and an arc between two variables represents a dependency. The strength of each dependency, as well as the uncertainty associated with these, is captured using probabilities and statistical distributions. When observed data are entered into the model for specific variables all the probabilities for, as yet unknown, unobserved or latent variables, are updated by Bayesian inference.

The BN model used for our analysis is shown in Figure 1 for the joint probability density function below:

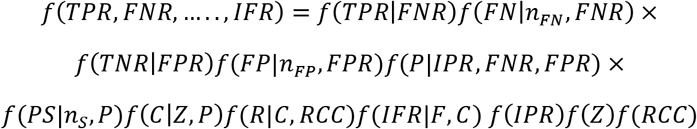

**Figure 1:**
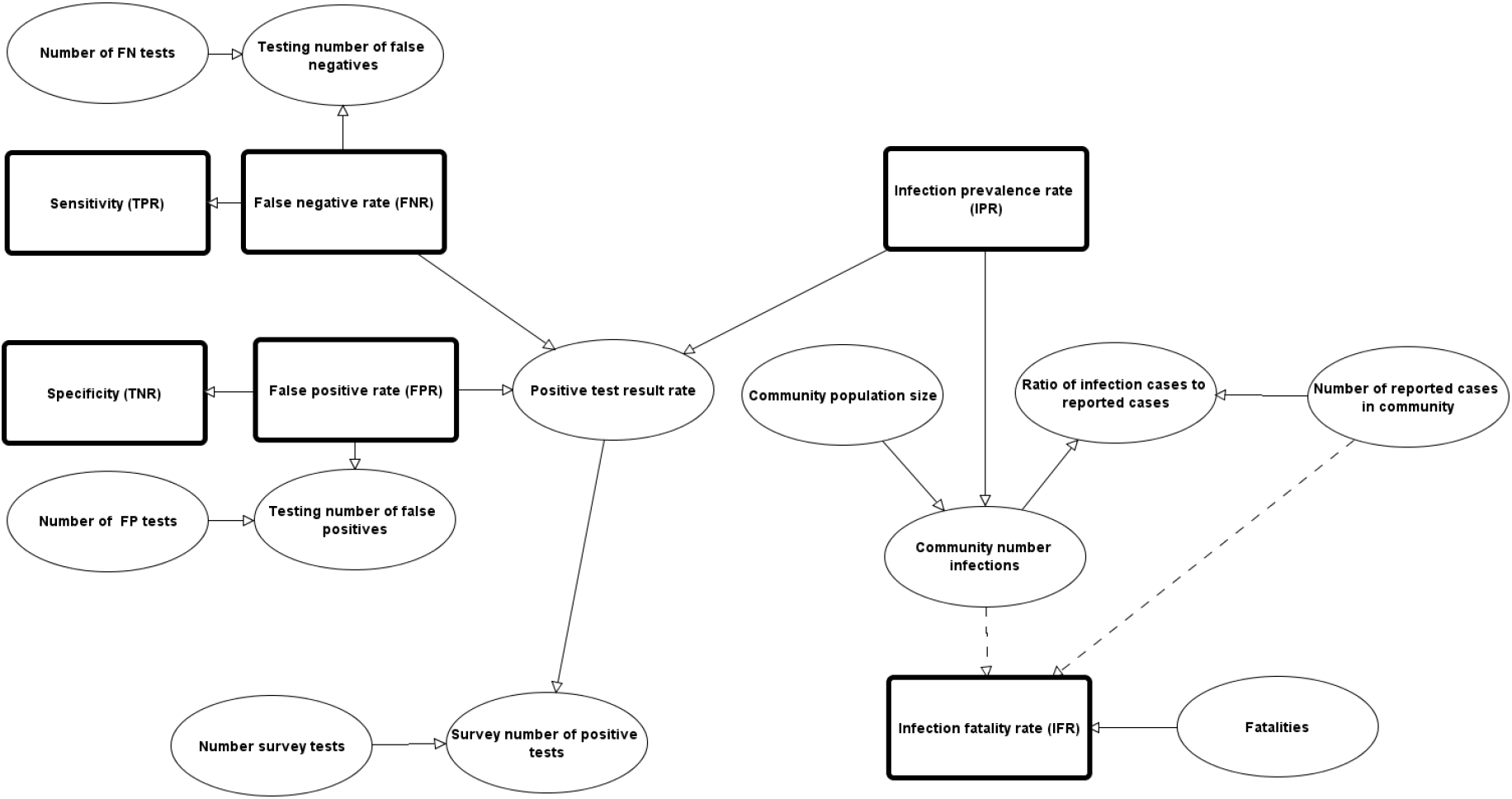
Bayesian Network Model to estimate Covid-19 IPR and IFR

This represents the conditonal and prior probability functions for these variables, as shown in Table 2. The statistical distributions and conditional probability densities are given in the appendix.

**Table 2:** Variables in Bayesian Network model

|  |  |  |  |
| --- | --- | --- | --- |
| $TPR$ | True positive rate (sensitivity) | $IPR$ | Infection Prevalance Rate |
| $TNR$ | True negative rate (specificity) | $PS$ | Survey number of positive tests |
| $FN$ | False negatives in testing | $n_s$ | Number of survey tests |
| $FP$ | False positives in testing | $IFR$ | Infection fatality rate |
| $n_{FN}$ | Number of false negative tests | $C$ | Community number of infections |
| $n_{FP}$ | Number of false positive tests | $Z$ | Community population size |
| $FPR$ | False positive rate | $R$ | Ratio of infection cases to reported cases |
| $FNR$ | False negatve rate | $RCC$ | Number of reported cases in community |
| $P$ | Positive test result rate | $F$ | Number of fatalilites |

Bayesian inference is performed in AgenaRisk [Agena 2020] which uses the dynamic discretization algorithm [Neil et al 2007].

The goal of a mass serological antibody survey is to estimate the infection prevalence rate *(IPR)* of the virus in the population. If we knew the *IPR*, the false negative rate *(FNR)*, and false positive rate *(FPR)* then we can determine the probability, *P*, of a positive test result as:

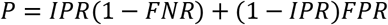

This is because there are two ways to test positive. The first is where someone is infected (probability *IPR)* and correctly test positive (probability (1 − *FNR))*, giving (1 − *FNR) × IPR*. The second is where someone is *not* infected, (1 − *IPR)*, but they falsely test positive, *FPR* and this gives us (1 − *IPR)FPR*.

Here we use the BN to solve the ‘inverse problem’, where we estimate the *IPR* given information about *FNR, FPR* and *P*, i.e. the test accuracy (sensitivity and specificity) and the number of positive tests from a given serological survey. Again, poor test accuracy will lead to poorer estimates of the *IPR* and, again, the size of the survey, in terms of samples taken, will determine the confidence in our estimates, with bigger samples translating to better information.

Once the BN model estimates the distribution for *IPR* this is used to estimate community infections, *C*, and ultimately the infection fatality rate, *IFR*.

## 5. Results using available data

### 5.1 Infection prevalence rates

If the serological testing surveys are imperfect what effect does this have on our estimated infection prevalence rates *(IPR)?*

To date very few serological surveys have been carried out and where they have been, some results have been difficult to obtain given the secrecy involved. The survey data and sources are listed below in Table 3. Note that here we are using raw data unadjusted for demographic or other population features.

**Table 3:**
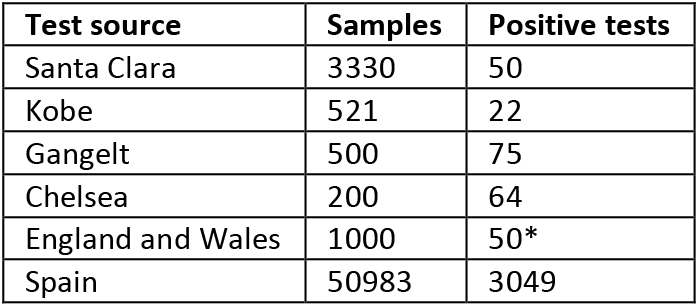
Serological surveys

| Test source | Samples | Positive tests |
| --- | --- | --- |
| Santa Clara | 3330 | 50 |
| Kobe | 521 | 22 |
| Gangelt | 500 | 75 |
| Chelsea | 200 | 64 |
| England and Wales | 1000 | 50* |
| Spain | 50983 | 3049 |

Note that the positive test results for England and Wales is inferred from public announcements made by the UK Government’s Chief Medical Officer [Blanchard 2020] about the UK ONS antibody survey [ONS 2020b].

We have the serological test results from the Santa Clara County, California USA, study [Biomedomics 2020] and Gangelt, Germany [FDA 2020]. However, for the Kobe, Japan, study we only had specificity and sensitivity point values and total sample sizes from Kurano [Karubo 2020] and had to infer the likely values. Also, we do not know what serological tests were applied in England and Wales. The UK government have, however, announced they were choosing Roche as one of their serological antibody test suppliers, and so we assume a test with equivalent accuracy was used [FDA 2020].

The full probability distributions that the BN model computes *IPR* for all cases are shown in Figure 2 and the associated summary statistics are listed in Table 4.

**Figure 2:**
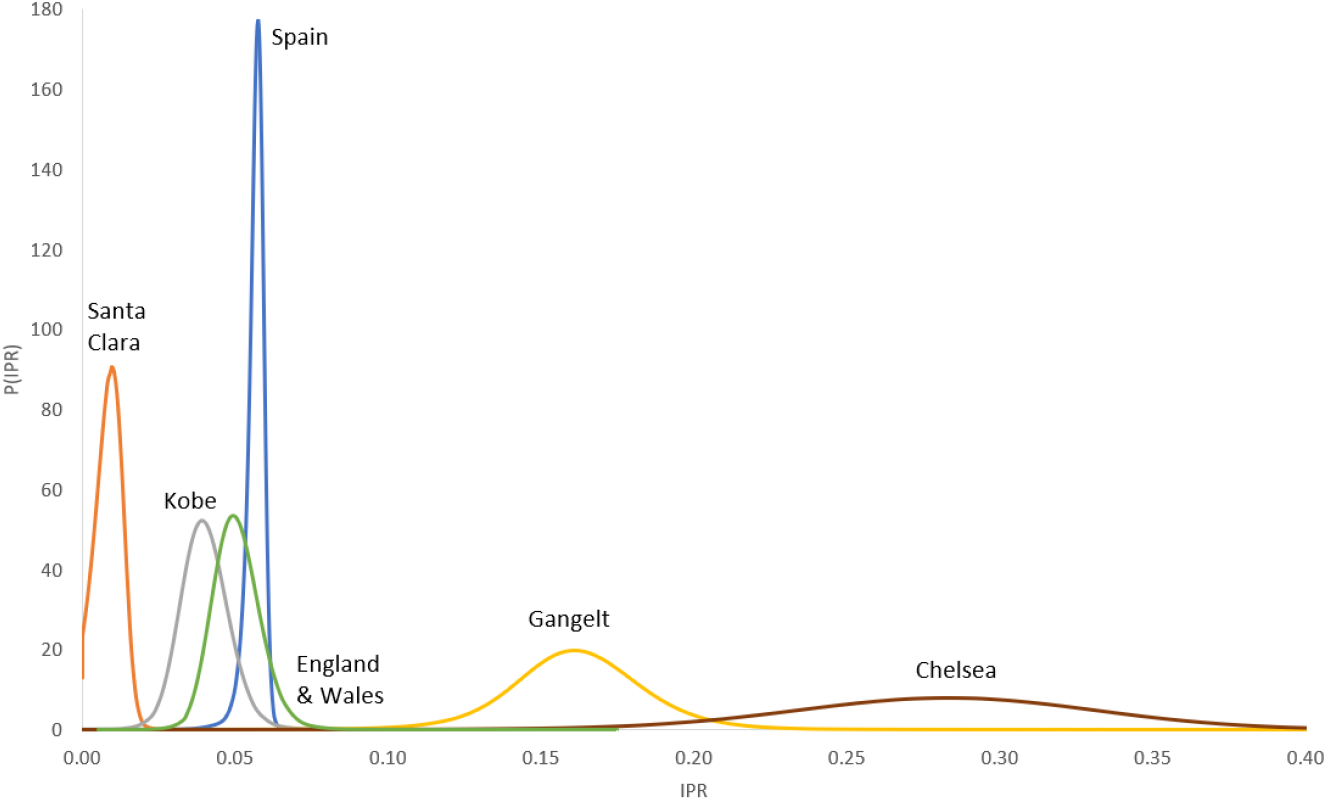
Posterior marginal distribution of Infection prevalence rate *(IPR)* for country/region cases

**Table 4:** Summary statistics for *IPR*

| Case | Test source | Assumption | mean | 95% CI |
| --- | --- | --- | --- | --- |
| Santa Clara | Premier Biotech | Actual source | 0.00879 (0.88%) | (0.0009, 0.0165) |
| Kobe | Kurabo | Actual source | 0.040 (0.4%) | (0.0254, 0.0562) |
| Gangelt | EUROIMMUN | Actual source | 0.162 (16.2%) | (0.1162, 0.0208) |
| Chelsea Mass. | BioMedomics | Actual source | 0.28 (28.1%) | (0.1795, 0.3780) |
| England and Wales | Roche | Hypothesised source | 0.0506 (0.51%) | (0.0368, 0.0670) |
| Spain | Orient Gene Biotech | Actual source | 0.05663 (0.56%) | (0.0496, 0.0617) |

From Figure 2 and Table 4 we can see that Chelsea Mass. USA and Gangelt Germany have relatively higher infection prevalence rates than Santa Clara USA, Kobe, Japan and England and Wales.

### 5.2 Estimates of community infections

Given our prevalence estimates, how does this compare to the case infection numbers? (those who test positive for Covid-19) i.e. how much higher is actual community infection compared to reported infection?

Now we have an estimate of the *IPR* we can use this to estimate the number of community infections, *C*, in a region or country, given the relevant community population size. We can also compare this estimate with the number of reported cases of Covid-19 in the community, to determine the extent to which Covid-19 is more or less widespread than thought and also to determine proportion of cases likely to be symptomatic/asymptomatic and severe/slight.

Source data on the actual number of Covid-19 infection cases have been taken from the relevant research papers or other sources, and in the case of the UK from worldometers, for mid-April, where it reported to be circa. 100,000 [Worldometers 2020].

The summary statistics form the distributions calculated from the BN of the number of community infections are given in Table 5.

**Table 5:**
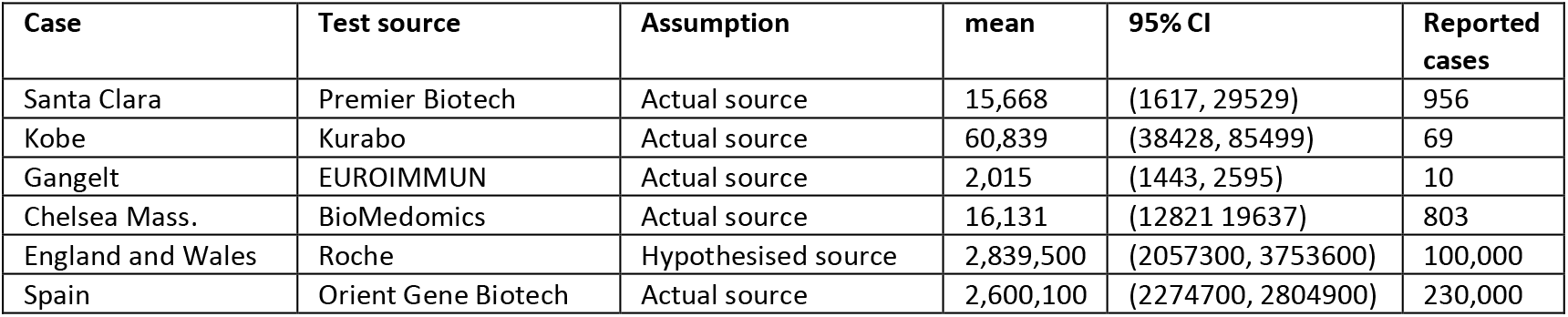
Summary statistics for Community number infections, *C*

| Case | Test source | Assumption | mean | 95% CI | Reported cases |
| --- | --- | --- | --- | --- | --- |
| Santa Clara | Premier Biotech | Actual source | 15,668 | (1617, 29529) | 956 |
| Kobe | Kurabo | Actual source | 60,839 | (38428, 85499) | 69 |
| Gangelt | EUROIMMUN | Actual source | 2,015 | (1443, 2595) | 10 |
| Chelsea Mass. | BioMedomics | Actual source | 16,131 | (12821, 19637) | 803 |
| England and Wales | Roche | Hypothesised source | 2,839,500 | (2057300, 3753600) | 100,000 |
| Spain | Orient Gene Biotech | Actual source | 2,600,100 | (2274700, 2804900) | 230,000 |

Table 5 shows that the estimated community prevalence is much higher than reported prevalence. Figure 4 and Table 6 show the results from the model of the ratio of estimated to reported cases and the effect that unreliability in survey testing has on the result. Clearly in all cases the extent of community prevalence is much higher than reported. There is however considerable uncertainty about the ratio distribution for Kobe and Santa Clara, given their confidence interval is so wide.

**Table 6:** Summary statistics for ratio of estimated community infections, *R*, to reported cases

| Case | Test source | Assumption | mean | 95% CI |
| --- | --- | --- | --- | --- |
| Santa Clara | Premier Biotech | Actual source | 16 | (1.68, 30.98) |
| Kobe | Kurabo | Actual source | 881 | (556.93, 1240.5) |
| Gangelt | EUROIMMUN | Actual source | 201 | (144.24, 259.7) |
| Chelsea Mass. | BioMedomics | Actual source | 14 | (8.95, 18.92) |
| England and Wales | Roche | Hypothesised source | 28 | (20.3, 37.5) |
| Spain | Orient Gene Biotech | Actual source | 11 | (9.88, 12.19) |

**Figure 3:**
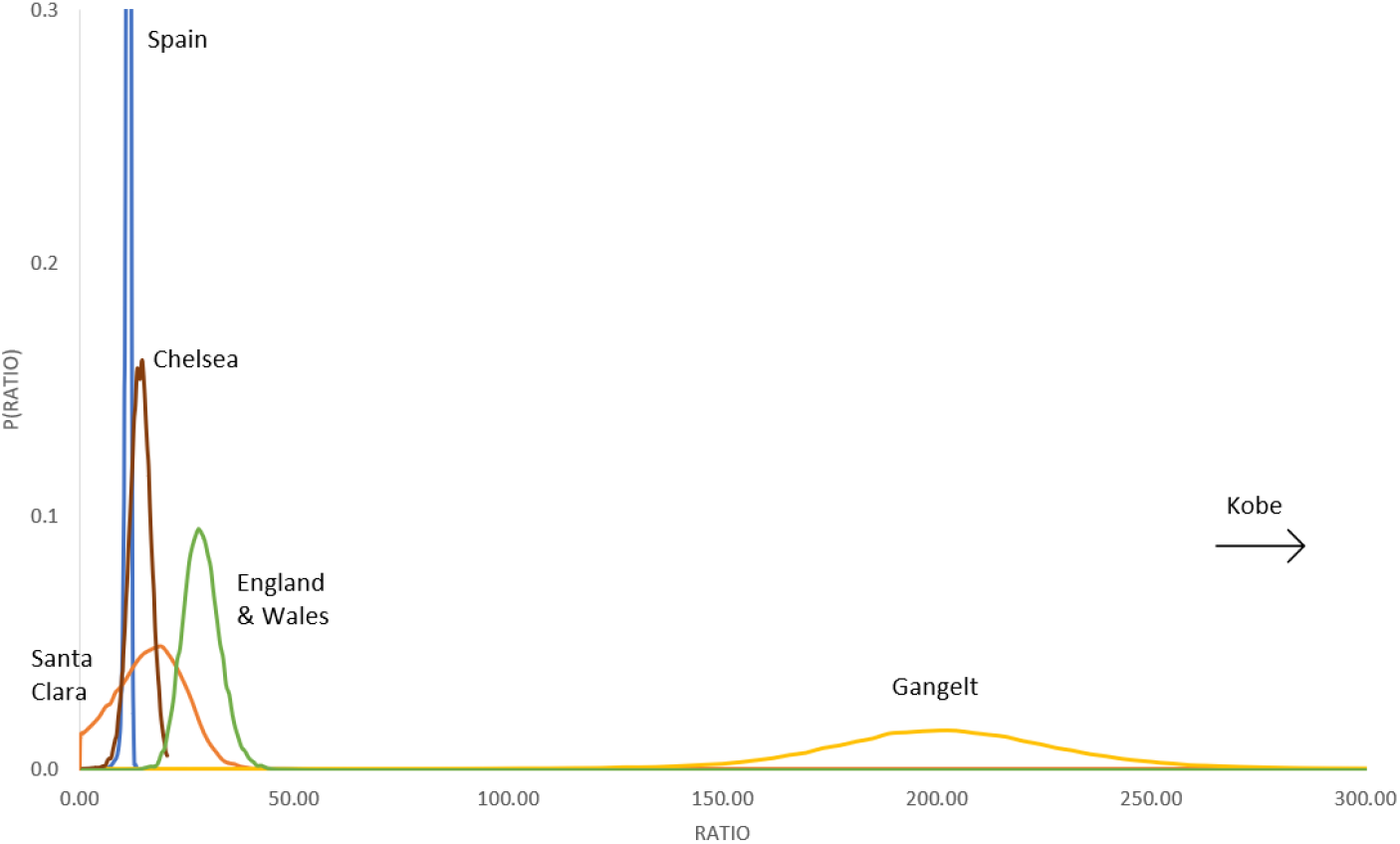
Posterior marginal distribution of Ratio of estimated community infections *(R)* to cases to reported cases by country/region cases (excluding Kobe, Japan)

### 5.3 Infection fatality rates

From reported fatality statistics what is the Covid-19 infection fatality rate *(IFR)?*

The final piece of the jigsaw puzzle is our estimation of *IFR*, for each of the countries and regions where we have data available, and here this includes, England and Wales and Chelsea Mass. USA, Santa Clara USA, Kobe Japan, Spain, and Gangelt, Germany.

The reported fatalities up to mid-April in the UK is 23,554 [ONS 2020b]. The reported fatality count for Chelsea, Massachusetts is 39 [Saltzman 2020] and for Santa Clara, California is 94 [Ioannidis 2020]. For Gangelt, Germany it is 7 [Streek et al 2020]. For Spain, the fatality count is 27,888 [Worldometers 2020] and for Kobe Japan it is 10 [Ioannidis 2020]. Figure 4 shows the predicted distributions from the BN model for each *IFR*, while Table 7 shows the summary statistics.

**Figure 4:**
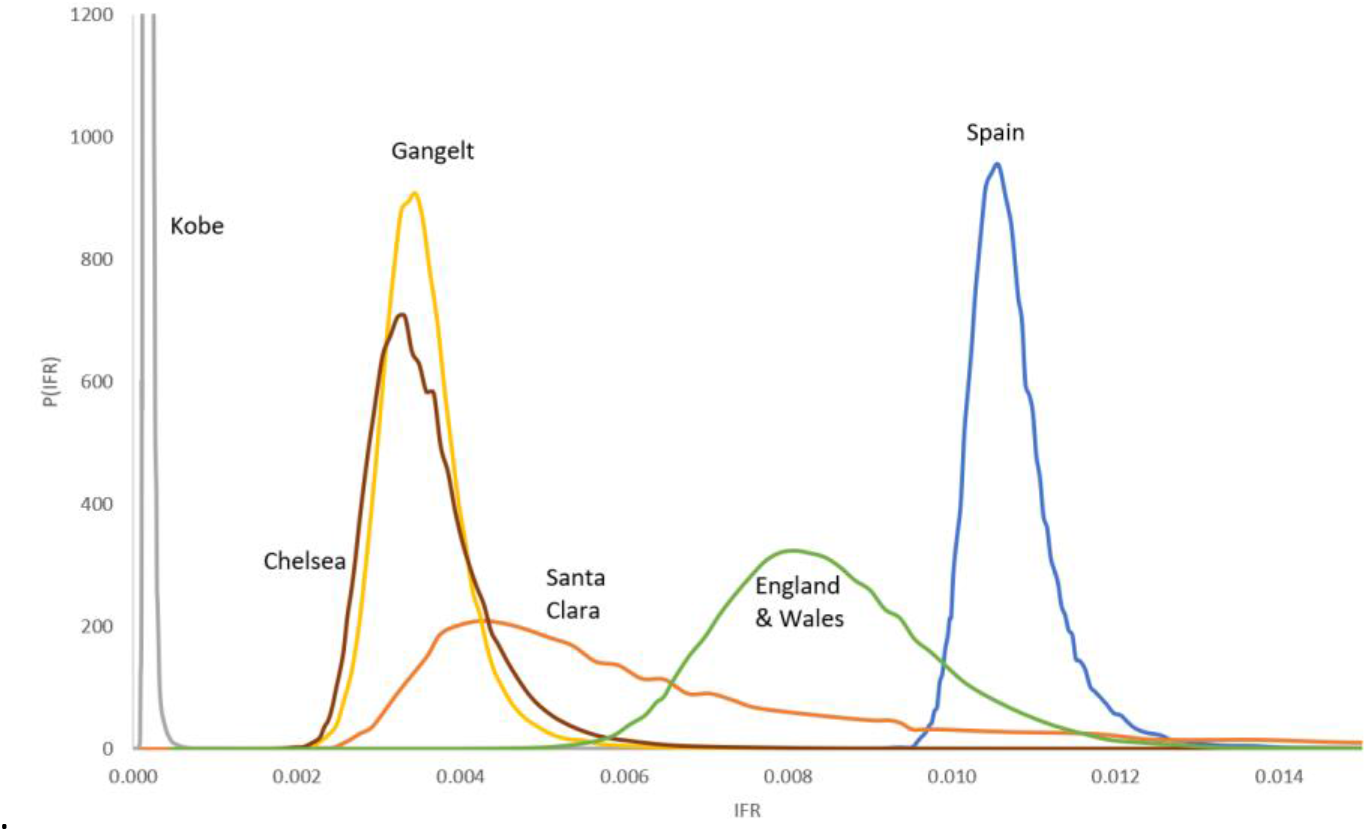
Posterior marginal distribution of Infection fatality rate *(IFR)* for country/region cases

**Table 7:** Summary statistics for the *IFR* rate

| Case | Test source | Fatality count | mode | mean | 95% CI |
| --- | --- | --- | --- | --- | --- |
| Santa Clara | Premier Biotech | 94 | 0.004 (0.4%) | 0.0102 (1.02%) | (0.0031, 0.0595) |
| Kobe, Japan | Kurabo | 10 | 0.00015 (0.001%) | 1.7271E-4 (0.0002%) | (1.16E-4, 2.60E-4) |
| Chelsea Mass. | BioMedomics | 39 | 0.003 (0.3%) | 0.0029 (0.29%) | (0.0025, 0.0054) |
| Gangelt, Germany | EUROIMMUN | 7 | 0.003 (0.3%) | 0.0036 (0.36%) | (0.0026, 0.0048) |
| England and Wales | Roche | 23,554 | 0.008 (0.8%) | 0.0085 (0.85%) | (0.0062, 0.0114) |
| Spain | Orient Gene Biotech | 27,888 | 0.010 (1.0%) | 0.0107 (0.107%) | (0.0099, 0.0122) |

The most likely (modal) *IFR* values are grouped for Santa Clara and Chelsea, both in the USA, with mode values of 0.3-0.4%. However, for Santa Clara there is higher uncertainty giving a long tail in the data hence the mean value is significantly higher at 1.02%. This shows the estimate for Santa Clara is less informative. Kobe, Japan is very unusual in comparison with the others with values an order of magnitude less at, 0.001%. The *IFR* for Spain has a low variance, which is unsurprising given the very large survey conducted, with a mean and mode rate close to each other centred around 1%. England and Wales lie between Spain and the USA/German values with a mode and mean of 0.8%.

### 5.5 Sensitivity to uncertainty

How does serological test quality affect the reliability of the above estimates of the infection fatality rate *(IFR)?* Likewise, how does uncertainty about fatality counts affect these estimates?

The preceding analysis has simply used the available ‘raw’ data without any adjustment for demographics or to take account of differences between countries and regions. In this section we choose two sources of uncertainty that have not been covered by other researchers and which strongly determine the sensitivity of results, especially for *IFR*. These concern fatality counts and the quality of the serological surveys themselves.

There is some controversy over UK fatality counts [Fenton et al 2020] and there appear to be many reasons not to trust the fatality count, including:

- Ambiguity and confusion about diagnostic criteria for Covid-19
- Care home deaths not certified by a qualified medical practitioner
- Hospital and other deaths signed off as “caused by” Covid-19 when they are “with” Covid-19
- The number of excess deaths could be much higher because of cases that remain undiagnosed

Similar concerns apply to Spain and potentially elsewhere. Given that these uncertainties work in both directions we must be careful to include under and over-estimates and not chose one to suit our prejudices or political outlook.

The Santa Clara study has been heavily criticised by some statisticians in a popular blog [Stats Blog 2020], mainly because subjects were recruited using Facebook and perceived issues with sample sizes and other aspects of the analysis undertaken. Similar observations and comments have been made about the Chelsea study because subjects were chosen for convenience and availability rather than at random.

Typically, experimenters would design their study to address the sensitivity of the results to uncertainties, using relevant and, potentially, causal classifications such as demographic stratification of the relevant population. However, given that we have observational rather than experimental data we can only perform a post hoc evaluation of the sensitivity of the results. Hence, in this section we address these major uncertainties, using our BN model. The lack of controls in these observational studies translates into less informative estimates and more variability.

We use the notion of ‘virtual’ evidence to assess how different observations might affect our results. In this way we view the observations not as ‘facts’ but as random variables, bounded by possible ranges of value. This enables us to ask hypothetical questions such as: What would the results have been had fatalities been exaggerated or the number of positives from a study been fewer?

For the Santa Clara *PS_obs_ = Binomial (n_s_,P)* = 50, meaning we have 50 positive results from the binomial model. Virtual evidence allows us to place a likelihood over the distribution for *PS_obs_* that reflects our uncertainty about the study design. Here we use *PS_obs_~Uniform(* 1,50) representing a sceptical belief that any value between one and fifty is equally likely from this study because the study design was such that it made it more likely that prior infected subjects would volunteer. Once we enter this virtual evidence into the Bayesian Network it will compute the posterior probability distribution using this likelihood instead of a single point observation. Bayesian inference then computes the posterior distribution using this ‘sceptical’ likelihood along with other evidence in the model. Clearly, the resulting posterior will not necessarily match the likelihood provided, and indeed may contradict it, perhaps demonstrating the scepticism was unwarranted. We can apply the same process to the fatality counts and apply ‘virtual evidence’ that reflects deeper uncertainties about whether people ‘died because of’ Covid-19 as opposed to ‘with’ Covid-19.

We applied virtual evidence in four cases, as shown in Table 8. Uncertainties about the study design are represented for Santa Carla and Chelsea, Mass., USA. Uncertainties about the true fatality count are represented for England & Wales and Spain.

**Table 8:**
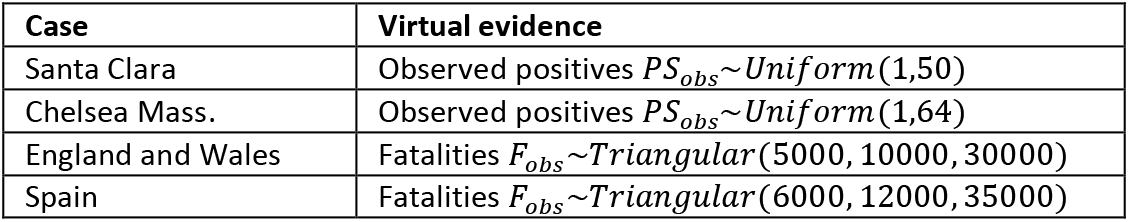
Virtual evidence scenarios

| Case | Virtual evidence |
| --- | --- |
| Santa Clara | Observed positives $PS_{obs} \sim \text{Uniform}(1, 50)$ |
| Chelsea Mass. | Observed positives $PS_{obs} \sim \text{Uniform}(1, 64)$ |
| England and Wales | Fatalities $F_{obs} \sim \text{Triangular}(5000, 10000, 30000)$ |
| Spain | Fatalities $F_{obs} \sim \text{Triangular}(6000, 12000, 35000)$ |

The sensitivity of *IFR* to these changes to the evidence are shown with the full distributions in Figure 5 and summary statistics in Table 9.

**Figure 5:**
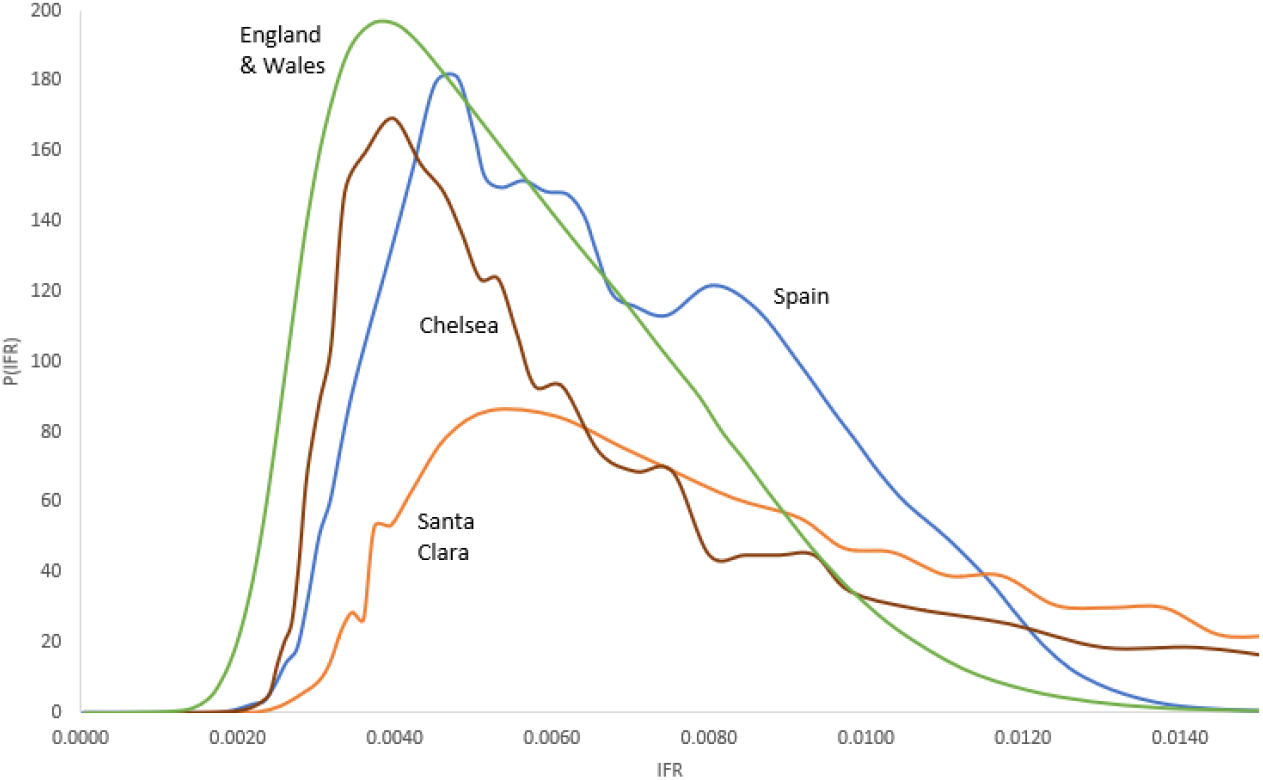
Posterior marginal distribution of Infection fatality rate *(IFR)* for country/region cases in virtual evidence scenarios

**Table 9:** Summary statistics for *IFR* in virtual evidence scenarios

| Case | Test source | Fatality count | mode | mean | 95% CI |
| --- | --- | --- | --- | --- | --- |
| Santa Clara | Premier Biotech | 94 | 0.005 (0.5%) | 0.0217 (2.17%) | (0.0038, 0.0968) |
| Chelsea Mass. | BioMedomics | 39 | 0.003 (0.3%) | 0.0124 (1.24%) | (0.0030, 0.0485) |
| England and Wales | Roche | 23,554 | 0.004 (0.4%) | 0.0056 (0.56%) | (0.0024, 0.0106) |
| Spain | Orient Gene Biotech | 27,888 | 0.004 (0.4%) | 0.0107 (0.107%) | (0.0032, 0.0118) |

The effect of virtual evidence is to increase the variability of the results, as one would expect. For the Santa Clara and Chelsea studies where we adjusted the observed survey positives the mean fatality rate estimates increase. However, the modal values do not change much at all, remaining around 0.3%-0.5%, suggesting that the BN model is balancing the sceptical explanations against other evidence in the model and discounting any virtual evidence that is perhaps too sceptical. Figure 6 shows the posterior marginal distributions that result from the application of virtual evidence on the number of positive tests for these studies. Clearly any strong scepticism that the number of genuine positives must be close to zero is not warranted and in the case of Chelsea observations less than 30 are difficult to justify. The model clearly ‘believes’ the survey results are credible.

**Figure 6:**
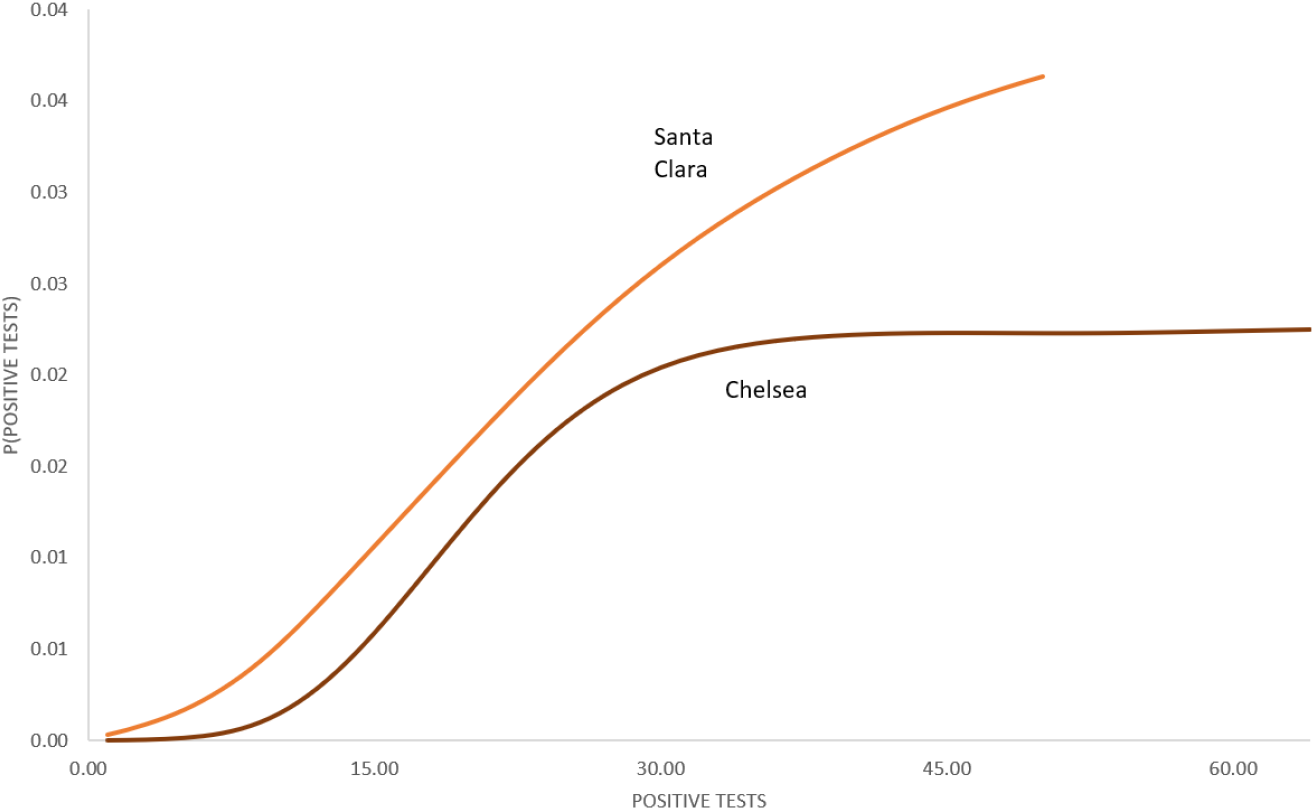
Posterior marginal distribution of Infection for the number of positive tests for the Santa Clara and Chelsea cases.

After the adjustment, distributions of the *IFR* across the USA, Spain and the UK look very similar; the most likely range for *IFR* is between 0.2% to 1%, with values beyond this looking less likely.

### 5.6 Comparison with influenza

How do our estimates of covid-19 *IFR* compare to influenza *IFR?* For comparison, the fatalities in England and Wales for the ‘higher than seasonal average’ in the year 1999/2000 were 21,290 excess deaths attributed to influenza like illness [Crofts et al 2002], from an infected population of 440,440 (influenza like illness reports) [ONS 2015c]. With a population of approximately 52m this is an *IPR* of just over 0.8% and a case fatality rate *CFR* of 4.8%. However, it should be noted that the equivalent of population level serological antibody testing is not routinely performed for influenza hence the *IPR* is likely to be underestimated and the *IFR* significantly less than the *CFR*.

## 6. Conclusion

Widely reported statistics on Covid-19 across the globe fail to take account of both the uncertainty of the data and possible explanations for this uncertainty. In this paper we have used a Bayesian Network (BN) model to estimate the Covid-19 *infection prevalence rate (IPR)* and *infection fatality rate (IFR)* for different countries and regions, where relevant data are available. This combines multiple sources of data in a single model.

We used this model, and available data on serological surveys, fatality counts and test accuracy, to answer the following questions: What is the accuracy of serological antibody tests under development and will it be sufficient to estimate population infection prevalence? What serological testing surveys have been done and what does the data tell us about the prevalence of community infection in different locations? If the serological testing surveys are imperfect what effect does this have on our estimated infection prevalence rates *(IPR)?* Given, our prevalence estimates, how does this compare to the case infection numbers? From reported fatality statistics what is the infection fatality rate *(IFR)?* How does serological test quality affect the reliability of these estimates? Likewise, how does uncertainty about fatality counts affect these estimates? How do our estimates of covid-19 *IFR* compare to influenza *IFR?*

The results show that Chelsea Mass. USA and Gangelt Germany have relatively higher infection prevalence rates *(IPR)* than Santa Clara USA, Kobe, Japan and England and Wales. In all cases the infection prevalence is significantly higher than has been widely reported, with much higher community infection rates in all locations. For Santa Clara and Chelsea, both in the USA, with most likely *IFR* values of 0.3-0.4%. Kobe, Japan is very unusual in comparison with the others with values an order of magnitude less than the others at, 0.001%. The *IFR* for Spain has a low variance, which is unsurprising given the large survey conducted, with an *IFR* centred around 1%. England and Wales lie between Spain and the USA/German values with an *IFR* around 0.8%. There remains some uncertainty around these estimates but an *IFR* greater than 1% looks remote. We used a Bayesian technique called ‘virtual evidence’ to test the sensitivity of the *IFR* to two significant sources of uncertainty: survey quality and uncertainty about death rates. In response the adjusted estimates for *IFR* that are most likely to be in the range 0.3%-0.5%. The unadjusted and adjusted Covid-19 *IFRs* calculated may be significantly less than that from the 1999/2000 UK influenza season, which had an *IPR* of just over 0.8% and a case fatality rate *CFR* of 4.8%. However, the influenza *IPR* is likely to be underestimated and the *IFR* significantly less than the *CFR* due to the absence of routine serological surveillance.

It is worth emphasising the significant problems facing any researcher with objectives like ours. There is a severe lack of transparency and important data sets are inaccessible. To support open debate these should be treated as a public good and shared openly throughout the scientific community in a timely manner.

## Data Availability

No primary data collected.

## Acknowledgements

This work was supported in part by the EPSRC under project EP/P009964/1: PAMBAYESIAN: Patient Managed decision-support using Bayesian Networks. We are grateful to Elma Neil for producing the template for7 the statistical graphs.

## Appendix – Bayesian Network Model

*f(TPR,FNR,…..,IFR) = f(TPRlFNR)f(FN|n_FN_,FNR) ×*

*f(TNR|FPR)f(FP|n_FP_,FPR)f(P|IPR, FNR, FPR) × f(PS|n_S_,P)f(C|Z,P)f(R|C,RCC)f(IFR|F, C) f(IPR)f(Z)f(RCC)*

*False Positive Rate: FPR~Uniform*(0,1)

*False Negative Rate: FNR~Uniform*(0,1)

*Specificity (FPR)* = 1 − *FPR*

*Sensitivity (TPR)* = 1 − *FNR*

*Testing number of false positives: FP~Binomial*(*n_FP_,FPR*)

*Number of FP tests: n_FP_*

*Testing number of false negatives: FN~Binomial*(*n_FN_,FNR*)

*Number of FN tests: n_FN_*

*Infection prevalence rate: IPR~Uniform*(0,1)

*Positive test result rate: P = IPR*(1 − *FNR*) + (1 − *IPR)FPR*

*Number of survey tests: n_S_*

*Survey number of positive tests: PS~Binomial*(*n_S_,P*)

*Community population size: Z~Uniform*(1,1*E*10)

*Number of reported Covid*19 *cases in the community: RCC*

*Community number of infections: C~max* (*RCC,Binomial*(*Z,IPR*))

*Ratio of infected cases to reported cases:* 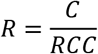

*Fatalities: F*

*Infection Fatality Rate: IFR = F/C*

Virtual evidence for England and Wales: *Fatalities: F_obs_~Triangular*(5000, 10000, 30000)

Virtual evidence for Spain: *Fatalities: F_obs_~Triangular*(6000, 12000, 35000)

Virtual evidence for Santa Clara, USA study: *PS_obs_~Uniform*(1,50)

Virtual evidence for Chelsea, USA study: *PS_obs_~Uniform*(1,64)

## Footnotes

1 Note that these rates can either be presented as percentages (ranging from 0-100%) or as probabilities (ranging from 0 to 1) as done in the formulas here.

